# Factors Associated with Measles Vaccine Drop-out Among Children Aged 12-23 Months in Selected Urban Slum of Dhaka

**DOI:** 10.64898/2025.12.18.25342636

**Authors:** Sayik Bin Alam, Md Foyjul Islam, Mehejabin Nurunnahar, Md. Tanvir Hossen, Md. Omar Qayum, Mohammad Rashedul Hassan, Tahmina Shirin

## Abstract

**Background:** Measles vaccine dropout between the first (MCV1) and second dose (MCV2) remains a public health concern in Bangladesh, particularly in urban slum settings. Identifying factors associated with vaccine dropout is essential to improve coverage and achieve the national measles elimination target by 2026.

**Objective:** To identify factors associated with measles vaccine dropout among children aged 12–23 months in an urban slum of Dhaka, Bangladesh.

**Methods:** A community-based case-control study was conducted in Korail slum, Dhaka, including 99 children (33 cases, 66 controls) selected purposively in 2023. Cases were children in Korail slum aged 12–23 months who received the first Pentavalent dose but missed MR1 or MR2; whereas the controls completed the measles vaccination schedule, matched by age and sex. Data on socio-demographic characteristics, healthcare access, and vaccination service experiences were collected using structured questionnaires. Bivariate and multivariate logistic regression analyses were performed to identify factors associated with vaccine dropout.

**Results:** Mean age of children was 16.2 months (SD ±2.6). Children of homemaker mothers had higher odds of dropout than those of working mothers (AOR: 4.84; 95% CI: 1.72–13.63). Second-born children were more likely to drop out compared to first-born (AOR: 5.3; 95% CI: 1.86– 15.34). Longer waiting times before vaccination (≥15 minutes) were reported by 75.86% of cases versus 59.32% of controls. Other factors, including maternal education and household size, were not significantly associated.

**Conclusion:** Maternal occupation, birth order, and service-related factors influence measles vaccine dropout in urban slums. Targeted interventions such as flexible vaccination schedules, reducing waiting times, and engaging caregivers of multiple children are essential to improve coverage and support Bangladesh’s measles elimination goal by 2026.

## Introduction

Measles is a highly contagious viral disease, particularly severe in young children and those with poor nutritional status, leading to complications such as bronchopneumonia, gastroenteritis, and otitis media. Immunization is one of the most cost-effective public health interventions, yet over 30 million infants in developing countries remain incompletely immunized each year[1]. Global measles mortality declined by 79%, from 546,800 deaths in 2000 to 114,900 in 2014, while in the South-East Asia Region incidence decreased from 70 to 16 cases per million population during the same period [2].In Bangladesh, confirmed measles cases decreased from 31.4 per million in 2019 to 1.34 in 2022 [3].

The World Health Organization (WHO) launched the Expanded Programme on Immunization (EPI) in 1974 to protect children from six major vaccine-preventable diseases, including measles [4]. Bangladesh adopted EPI in 1979 and rapidly expanded it nationwide, achieving significant success [4]. In 2013, the WHO South-East Asia Region set a goal for measles elimination by 2020 (Resolution SEA/RC66/R5) [2]. By 2016, regional MCV2 coverage reached 73%, and several countries have since eliminated measles and rubella [5].

The World Health Organization (WHO) recommends reducing vaccination dropout rates to less than 10% to ensure effective protection against vaccine-preventable diseases. However, measles vaccination dropout remains a major challenge in many low- and middle-income countries (LMICs). A recent study in Somalia found that more than 40% of children aged 15–23 months did not complete measles vaccination, with parental education, maternal occupation, family income, birth order, and negative attitudes as key factors [1]. Similarly, a systematic review in Ethiopia reported dropout rates above the WHO target, linked to poor antenatal care, home delivery, and limited access to health facilities [2].

Despite progress, inequities persist. The 2019 Coverage Evaluation Survey in Bangladesh found lower valid full vaccination coverage in urban areas (84.6%) compared to rural (89.5%).

The World Health Organization (WHO) recommends keeping vaccination dropout rates below 10% to ensure adequate protection against vaccine-preventable diseases[6]. However, measles vaccine dropout remains a significant challenge in low- and middle-income countries (LMICs) [7]. A recent study in Somalia found that more than 40% of children aged 15–23 months did not complete measles vaccination, with parental education, maternal occupation, family income, birth order, and negative attitudes as key factors [8]. A systematic review in Ethiopia also reported high dropout rates, linked to poor antenatal care, home delivery, and limited access to health facilities [2]. Another a case-control study in central Ethiopia found that lack of reminders during postnatal care, limited ANC visits, long waiting times, low maternal education, poor awareness of the second measles dose, and missed vitamin A supplementation significantly increased the likelihood of dropout[9].

In Bangladesh, Urban children had higher drop-out rates from Penta1–MR1 (5.5% vs. 3.4%) and MR1–MR2 (5.0% vs. 4.4%) [10]. Urban slum dwellers, estimated at 1.8 million people, face particular challenges, including poor access to antenatal care, institutional delivery, and immunization services [6,7]. Their frequent mobility and financial or social barriers further hinder continuity of care[11].

Given these disparities, strengthening immunization among slum populations is essential for achieving national and global targets, including the Sustainable Development Goals. This study aimed to identify socio-demographic factors, parental attitudes and perceptions, and maternal health-seeking behaviors associated with measles vaccine drop-out among children aged 12–23 months in a selected urban slum of Dhaka.

## Materials and Methods

### Study Design and Setting

A community-based case–control study was conducted from October 2023 to April 2024 in the Korail slum, one of the largest informal settlements under Dhaka North City Corporation (DNCC). The slum spans about 90 acres, houses over 50,000 residents, and has an annual vaccination target of approximately 1,750 children.

### Study Population and Sampling

The study population consisted of children aged 12–23 months and their parents or caregivers who had resided in Korail for at least six months. Cases were defined as children who had received the first dose of the Pentavalent vaccine but not either the first (MR1) or second (MR2) dose of the Measles–Rubella (MR) vaccine, verified through the EPI registration book. Controls were children who had received the Pentavalent vaccine and completed the age-appropriate MR vaccination schedule. Cases and controls were drawn from the same community and matched by age, sex. Exclusion criteria included incomplete or unverifiable vaccination records, severe illness or comorbidities in the child, or refusal of consent by caregivers.

Sample size was calculated using Epi Info StatCalc version 7.2 with a 95% confidence level, 80% power, and a 1:2 case-to-control ratio. Using an odds ratio of 3.57 for birth order, with 42.2% exposure among controls as reported in a previous study[12], the required sample was 99 children (33 cases and 66 controls). A purposive sampling strategy was applied. Eligible children were identified from EPI registers at Korail vaccination centers, and data collectors, assisted by local guides, visited households to recruit parents or caregivers until the desired sample size was achieved.

### Data Collection tools and procedure

Data were collected from 15/10/2023 to 30/04/2024 using a pre-tested, semi-structured questionnaire adapted from the National Coverage Evaluation Survey (CES) 2019. Face-to-face interviews were conducted with the parents or caregivers in Bangla after obtaining written informed consent. The questionnaire covered four domains: socio-demographic and parental characteristics (child’s sex, parental age, education, occupation, mother’s age at marriage and first birth, caregiver); family and household factors (family size, monthly expenditure, birth order, and birthplace of child); health service–related characteristics (antenatal and postnatal care visits, waiting time during vaccination, and post-vaccination complications); and parental attitudes and perceptions regarding measles disease, schedule, side effects.

Operational definitions were applied to ensure consistency. A “drop-out” was defined as failure to return for subsequent vaccine doses. “Penta1–MR1 drop-out” referred to children who received Penta1 but not MR1, while “MR1–MR2 drop-out” referred to those who received MR1 but not MR2. Parental attitudes were defined as beliefs and perceptions regarding the necessity, safety, and benefits of vaccination. Service delivery factors referred to accessibility, availability of vaccines and staff, quality of service, and health education. Maternal health-seeking behavior was defined as the use of healthcare services during pregnancy and postpartum that might influence child immunization.

Data collectors received training on study objectives, interview techniques, and informed consent procedures. The principal investigator reviewed completed questionnaires daily to ensure completeness and accuracy. Confidentiality was maintained throughout; hard copies were securely stored at IEDCR, and electronic data were de-identified and password protected.

### Statistical Analysis

Data entry was performed in Microsoft Excel, and analysis in STATA version 17. Means and standard deviations summarized continuous variables, and proportions described categorical variables. Chi-square or Fisher’s exact tests were used for bivariate analysis. Logistic regression was conducted to identify independent predictors of measles vaccine drop-out, with statistical significance set at *p* < 0.05.

### Ethical Considerations

Ethical approval for this study was obtained from the Institutional Review Board of the Institute of Epidemiology, Disease Control and Research (IEDCR). Permission was also granted by the Dhaka North City Corporation authorities. Written informed consent was obtained from all respondents. Confidentiality and anonymity were maintained by de-identifying data, securely storing hard copies at IEDCR, and password-protecting electronic files. Participation was voluntary, and caregivers were free to withdraw from the study at any time without any consequences.

## Results

A total of 99 children aged 12–23 months were included in this study, with 33 cases (drop-out) and 66 controls (non-drop-out).

### Socio-demographic and parental characteristics

Among the children, 54.6% were male and 45.5% were female, with no difference between cases and controls. Most mothers were aged 20–29 years in both groups (78.8% of cases and 68.2% of controls), while the majority of fathers were ≥30 years (57.6% of cases vs. 48.5% of controls). More mothers of cases were homemakers (63.6%) compared to controls (37.9%), while working mothers were more frequent among controls (62.1%). Regarding education, 40.0% of mothers of cases had education up to primary level compared to 31.8% among controls, while 69.7% of fathers of cases had secondary or higher education compared to 51.5% of controls. Early marriage (<18 years) was reported by 6.1% of case mothers and 15.2% of control mothers, whereas most gave their first birth at ≥18 years in both groups [Table 1].

**Table 1.**
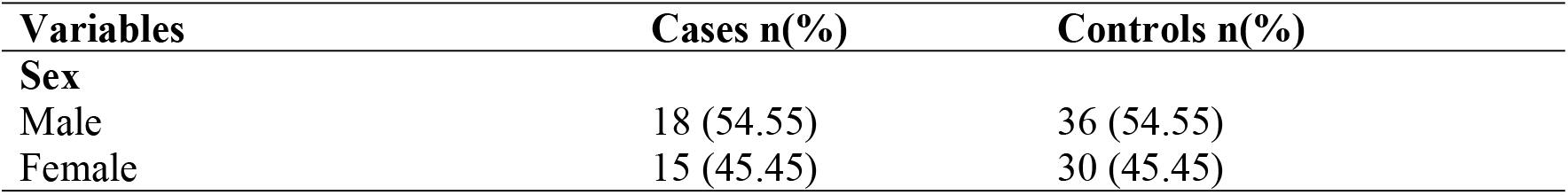
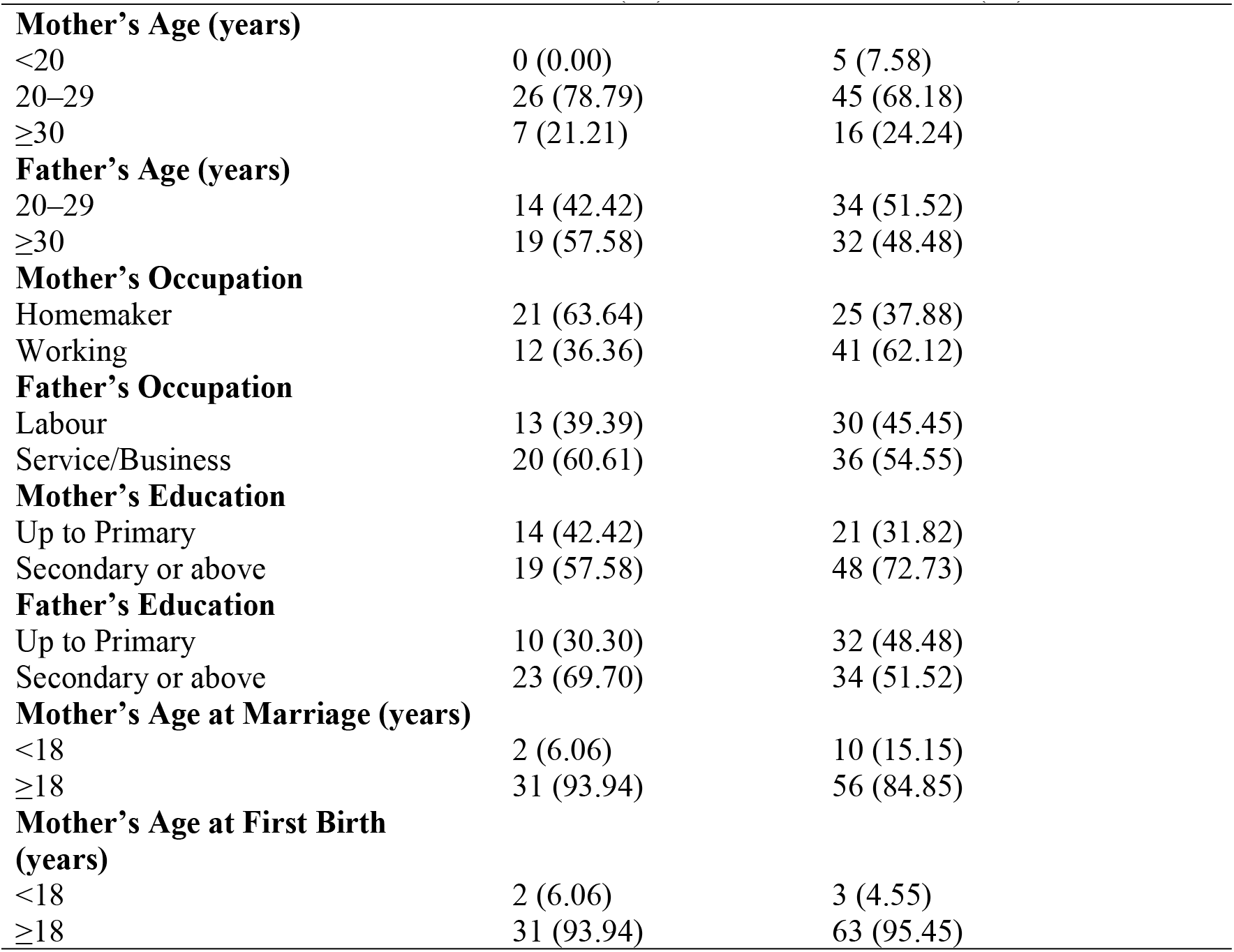
Socio-demographic and parental characteristics of cases and controls among children aged 12-23 months in Korail slum of Dhaka, 2023 (N=99)

| Variables | Cases n(%) | Controls n(%) |
| --- | --- | --- |
| **Sex** |  |  |
| Male | 18 (54.55) | 36 (54.55) |
| Female | 15 (45.45) | 30 (45.45) |

| **Variables** | **Cases n(%)** | **Controls n(%)** |
| --- | --- | --- |
| **Mother's Age (years)** |  |  |
| <20 | 0 (0.00) | 5 (7.58) |
| 20–29 | 26 (78.79) | 45 (68.18) |
| ≥30 | 7 (21.21) | 16 (24.24) |
| **Father's Age (years)** |  |  |
| 20–29 | 14 (42.42) | 34 (51.52) |
| ≥30 | 19 (57.58) | 32 (48.48) |
| **Mother's Occupation** |  |  |
| Homemaker | 21 (63.64) | 25 (37.88) |
| Working | 12 (36.36) | 41 (62.12) |
| **Father's Occupation** |  |  |
| Labour | 13 (39.39) | 30 (45.45) |
| Service/Business | 20 (60.61) | 36 (54.55) |
| **Mother's Education** |  |  |
| Up to Primary | 14 (42.42) | 21 (31.82) |
| Secondary or above | 19 (57.58) | 48 (72.73) |
| **Father's Education** |  |  |
| Up to Primary | 10 (30.30) | 32 (48.48) |
| Secondary or above | 23 (69.70) | 34 (51.52) |
| **Mother's Age at Marriage (years)** |  |  |
| <18 | 2 (6.06) | 10 (15.15) |
| ≥18 | 31 (93.94) | 56 (84.85) |
| **Mother's Age at First Birth (years)** |  |  |
| <18 | 2 (6.06) | 3 (4.55) |
| ≥18 | 31 (93.94) | 63 (95.45) |

### Family and household characteristics

Most children were cared for by their mothers in both groups (66.7% of cases vs. 51.5% of controls). The majority of families were small (≤5 members) in both cases (81.8%) and controls (83.3%). Household monthly expenditure was predominantly between 10,001–20,000 BDT (84.9% of cases and 72.7% of controls). Birth order differed notably between groups: second-born children were more frequent among cases (54.6%) than controls (27.3%), whereas first-borns were more common among controls (66.7%) than cases (42.4%). Most children were delivered at health facilities (60.6% of cases and 71.2% of controls), while home births with skilled attendants were more common among cases (30.3%) [Table 2].

**Table 2.** Family and household characteristics of cases and controls among children aged 12-23 months in Korail slum of Dhaka, 2023 (N=99)

| Variables | Cases n(%) | Controls n(%) |
| --- | --- | --- |
| **Caregiver of child** |  |  |
| Mother | 22 (66.67) | 34 (51.52) |
| Others | 11 (33.33) | 32 (48.48) |
| **Family size** |  |  |
| ≤5 | 27 (81.82) | 55 (83.33) |
| >5 | 6 (18.18) | 11 (16.67) |
| **Monthly expenditure (BDT)** |  |  |
| ≤10000 | 3 (9.09) | 5 (7.58) |
| 10001–20000 | 28 (84.85) | 48 (72.73) |
| ≥20000 | 2 (6.06) | 13 (19.70) |
| **Birth order** |  |  |
| First-born | 14 (42.42) | 44 (66.67) |
| Second-born | 18 (54.55) | 18 (27.27) |
| Third-born or higher | 1 (3.03) | 4 (6.06) |
| **Birthplace of child** |  |  |
| Home (TBA) | 3 (9.09) | 9 (13.64) |
| Home (SBA) | 10 (30.30) | 10 (15.15) |
| Health facility | 20 (60.61) | 47 (71.21) |

### Health service–related characteristics

More than half of both cases (51.5%) and controls (59.1%) reported ≥4 ANC visits. However, fewer cases (42.4%) than controls (24.2%) had ≥4 PNC visits. Waiting time during vaccination was ≥15 minutes for most cases (75.9%) compared to 59.3% of controls. Almost all mothers received TT vaccination (97.0% of cases and 100% of controls). Complications following vaccination were reported by 72.7% of cases and 62.1% of controls [Table 3].

**Table 3.** Health service–related characteristics of cases and controls among children aged 12-23 months in Korail slum of Dhaka, 2023 (N=99)

| Variables | Cases n(%) | Controls n(%) |
| --- | --- | --- |
| **ANC visits** |  |  |
| <4 | 16 (48.48) | 27 (40.91) |
| ≥4 | 17 (51.52) | 39 (59.09) |
| **PNC visits** |  |  |
| <4 | 19 (57.58) | 50 (75.76) |
| ≥4 | 14 (42.42) | 16 (24.24) |
| **Waiting time during vaccination** |  |  |
| **(Cases = 29, Controls = 59)** |  |  |
| <15 minutes | 7 (24.14) | 24 (40.68) |
| ≥15 minutes | 22 (75.86) | 35 (59.32) |
| **Mother received TT vaccine** |  |  |
| Yes | 32 (96.97) | 66 (100.00) |
| No | 1 (3.03) | 0 (0.00) |
| **Complications after vaccination** |  |  |
| Yes | 24 (72.73) | 41 (62.12) |
| No | 9 (27.27) | 25 (37.88) |

### Bivariate logistic regression analysis

In bivariate analysis, mother’s occupation and child’s birth order showed significant associations with measles vaccine drop-out. Homemaker mothers were more likely to have children with drop-out compared to working mothers (COR: 2.87, 95% CI: 1.21–6.82, *p*=0.017). Second-born children had higher odds of drop-out compared to first-borns (COR: 3.14, 95% CI: 1.29–7.63, *p*=0.011). Other factors, including parental education, family size, household expenditure, ANC/PNC visits, place of delivery, and caregiver type, did not show statistically significant associations [Table 4].

**Table 4.** Bivariate logistic regression of factors associated with measles vaccine drop-out among children aged 12-23 months in Korail slum of Dhaka, 2023 (N=99)

| Variables | Cases<br>n(%) | Controls<br>n(%) | COR(95%CI) | p-value |
| --- | --- | --- | --- | --- |
| **Mothers Age during Marriage** |  |  |  |  |
| <18 years | 2(16.67) | 10(83.33) |  |  |
| ≥18 years | 31(35.63) | 56(64.37) | 2.76(0.56-13.44) | 0.207 |
| **Mothers Age During at the time of First Birth** |  |  |  |  |
| <18 years | 2(40.0) | 3(60.0) |  |  |
| ≥18 years | 31(32.97) | 63(67.03) | 0.74(0.12-4.64) | 0.746 |
| **Academic Qualification of the Mother** |  |  |  |  |
| Up to Primary | 14(40.0) | 21(60.0) |  |  |
| Secondary or above | 19(28.36) | 48(71.64) | 0.63(0.26-1.5) | 0.3 |
| **Occupation of the Mother** |  |  |  |  |
| Working | 12(22.64) | 41(77.36) |  |  |
| Homemaker | 21(45.65) | 25(54.35) | 2.87(1.21-6.82) | 0.017 |
| **Academic Qualification of the Father** |  |  |  |  |
| Up to Primary | 10(30.30) | 32(48.48) |  |  |
| Secondary or above | 23(69.70) | 34(51.52) | 2.2(0.89-5.24) | 0.087 |
| **Occupation of the Father** |  |  |  |  |
| Labour | 13(30.23) | 30(69.77) |  |  |
| Service | 19(38.0) | 31(62.0) | 1.4(0.59-3.36) | 0.433 |
| Business | 1(16.67) | 5(83.33) | 0.46(0.04-4.35) | 0.499 |
| **Family Size** |  |  |  |  |
| ≤5 | 27(32.93) | 55(67.07) |  |  |

| **Variables** | **Cases**<br>**n(%)** | **Controls**<br>**n(%)** | **COR(95%CI)** | **p-value** |
| --- | --- | --- | --- | --- |
| >5 | 6(35.29) | 11(64.70) | 1.11(0.37-3.3) | 0.851 |
| **Monthly expenditure of the family** |  |  |  |  |
| ≤10000 | 3(37.5) | 5(62.5) |  |  |
| 10001-20000 | 28(36.8) | 48(63.16) | 0.97(0.21-4.38) | 0.971 |
| ≥20000 | 2(13.33) | 13(86.67) | 0.25(0.03-2.02) | 0.196 |
| **Birth Place of the Child** |  |  |  |  |
| Home | 13(40.62) | 19(59.37) |  |  |
| Health Facility | 20(29.85) | 47(70.15) | 0.62(0.25-1.49) | 0.289 |
| **Sex of the Child** |  |  |  |  |
| Male | 18(33.33) | 36(66.67) |  |  |
| female | 15(33.33) | 30(66.67) | 1(0.43-2.3) | 1.00 |
| **Caregiver of child** |  |  |  |  |
| Mother | 22 (66.67) | 34 (51.52) |  |  |
| Others | 11 (33.33) | 32 (48.48) | 0.53(0.22-1.26) | 0.154 |
| **Birth Order of the Child** |  |  |  |  |
| First-born | 14(24.14) | 44(75.86) |  |  |
| Second-born | 18(50) | 18(50) | 3.14(1.29-7.63) | 0.011 |
| Third-born or higher | 1(20) | 4(80) | 0.78(0.08-7.62) | 0.154 |
| **Number of ANC visit** |  |  |  |  |
| 4 visits | 16(37.2) | 27(62.8) |  |  |
| ≥ 4 visits | 17(30.35) | 39(69.65) | 0.74(0.31-1.70) | 0.474 |
| **Number of PNC visit** |  |  |  |  |
| 4 visits | 19(27.5) | 50(72.5) |  |  |
| ≥ 4 visits | 14(46.67) | 16 (53.33) | 2.3(0.94-5.60) | 0.066 |
| **Have to wait long time during vaccination** |  |  |  |  |
| Never | 3(30) | 7(70) |  |  |
| Sometimes | 27(46.55) | 31(53.45) | 2.03(0.48-8.64) | 0.002 |
| Always | 3(9.68) | 28(90.32) | 0.25(0.04-8.64) | 0.132 |
| **Developed complications after vaccination** |  |  |  |  |
| Yes | 24(36.92) | 41(63.08) |  |  |
| No | 9(26.47) | 25(73.53) | 0.61(0.246-1.53) | 0.246 |

### Multivariate logistic regression analysis

After adjusting for confounders, mother’s occupation and birth order remained significantly associated with measles vaccine drop-out. Children of homemaker mothers had 4.84 times higher odds of drop-out compared to children of working mothers (AOR: 4.84, 95% CI: 1.72–13.63, *p*=0.003). Similarly, second-born children were 5.3 times more likely to drop out compared to first-borns (AOR: 5.3, 95% CI: 1.86–15.34, *p*=0.002). No significant association was observed for third-born or higher-order children [Table 5].

**Table 5.** Multivariate logistic regression of factors associated with measles vaccine drop-out among children aged 12-23 months in Korail slum of Dhaka, 2023 (N=99)

| Variables | COR (95% CI) | AOR (95% CI) | p-value |
| --- | --- | --- | --- |
| **Mother's Occupation** |  |  |  |
| Working | Ref | Ref |  |
| Homemaker | 2.87 (1.21–6.82) | 4.84 (1.72–13.63) | 0.003 |
| **Birth Order of the Child** |  |  |  |
| First-born | Ref | Ref |  |
| Second-born | 3.14 (1.29–7.63) | 5.3 (1.86–15.34) | 0.002 |
| Third-born or higher | 0.78 (0.08–7.62) | 0.54 (0.27–1.08) | 0.986 |

## Discussion

In this case-control study, we aimed to investigate the factors associated with measles vaccine dropout among children aged 12-23 months in the urban slums of Dhaka. Measles vaccine dropout remains a significant public health issue, particularly in socioeconomically disadvantaged communities like urban slums, where healthcare access and service quality are often inadequate.

This study in Dhaka’s urban slums identified several key factors associated with measles vaccine dropout. Notably, maternal occupation and family composition stood out. We found that children of homemaker mothers were significantly more likely to miss the second measles dose (adjusted odds ratio [AOR]≈4.8) than children of employed mothers. This suggests that employment outside the home may be associated with better healthcare-seeking behaviour, possibly due to greater awareness, access to healthcare services, decision making capability or financial stability. Similar findings have been reported in a study conducted in Nigeria, where employed mothers were more likely to ensure their children completed the vaccination schedule, possibly due to better exposure to health information or increased financial independence [13,14].

Furthermore, birth order was another factor significantly associated with vaccine dropout. Children who were second-born had higher odds of being in the dropout group (AOR: 5.3, 95% CI: 1.86-15.34), which aligns with findings from studies in Ethiopia and India, where higher birth order was linked to lower vaccination coverage[15]. Families with multiple children may face logistical challenges, such as the cost of traveling to vaccination centres or difficulties in managing various children’s healthcare needs, which increases the likelihood of dropout[16].

By contrast, maternal education was not significantly different between cases and controls in our data. However, previous studies have emphasized that maternal education is a crucial factor in child vaccination. For example, a study in Ethiopia discovered that children of mothers with higher education were more likely to be fully vaccinated compared to those whose mothers had no formal education[9,15]. Although our findings did not show a significant association, it is essential to consider the role of education in increasing awareness of the importance of completing the vaccination schedule[17].

The results of our study highlight the importance of healthcare access in vaccine adherence. Both cases and controls reported a high number of antenatal care (ANC) visits (≥4 visits for 51.52% of cases and 59.09% of controls), but postnatal care (PNC) visits were significantly less frequent among both cases and controls. A research from Zambia demonstrated that mothers who attended fewer PNC visits were less likely to complete their children’s vaccination schedule[18]. Maternal health-seeking behaviour can serve as an early indicator for potential measles vaccine dropouts. Studies have shown that factors such as limited antenatal care visits, lack of reminders during postnatal care, and prolonged waiting times at vaccination sites are associated with higher rates of measles vaccination dropout. For instance, mothers with fewer than two antenatal care visits were nearly five times more likely to have children who missed their second measles vaccine dose. Similarly, mothers who did not receive reminders during postnatal care were over five times more likely to have children who dropped out of the vaccination schedule. These findings suggest that monitoring maternal engagement with healthcare services can help identify children at risk of incomplete immunization[19].

Healthcare worker engagement also played a crucial role. Although most of both groups reported healthcare worker visited during pregnancy, the consistency and quality of these interactions could have influenced vaccine adherence. Strengthening follow-up systems and ensuring continuity of care during the postnatal period are crucial strategies for improving vaccine coverage[20].

The study found that with 42.42% of controls always had to wait long times compared to only 9.09% of cases (p = 0.002). Caregivers of controls might perceive vaccination as a high-priority activity, making them willing to tolerate long waiting times. Health education or awareness campaigns in the community might have contributed to this perception. For cases, other barriers, such as work responsibilities might outweigh the relatively shorter waiting times. It underscores that waiting time alone is not the sole determinant of vaccine adherence. Long waiting times have been identified as barriers to vaccine uptake in various settings. Again, A higher proportion of cases compared to controls experienced a waiting time of 15 minutes or more before vaccination, potentially contributing to vaccine dropout. A study in Uganda found that parents who faced long waiting times at healthcare facilities were less likely to return for follow-up vaccinations[21]. Improving service efficiency and reducing wait times in vaccination centres could have a significant impact on improving vaccine completion rates. To address this issue, the Expanded Program on Immunization (EPI) should implement efficient queue management strategies to enhance service delivery and reduce waiting times. This can include introducing dedicated vaccination time slots, allowing caregivers to plan their visits more effectively. Additionally, deploying extra healthcare staff during peak hours can help manage high patient flow and expedite the vaccination process. Furthermore, implementing pre-registration or appointment-based systems can help distribute the workload more evenly and minimize congestion at vaccination sites[22]. By optimizing these operational aspects, EPI can improve caregiver satisfaction, enhance vaccine coverage, and ultimately strengthen the immunization program’s effectiveness.

In terms of post-vaccination complications, a higher proportion of cases reported complications, compared to controls, Though the findings were not statistically significant. While most vaccine-related complications are minor, they can cause concern among parents, leading to vaccine hesitancy. This finding is consistent with a study conducted in Pakistan, where adverse effects following vaccination were a significant reason for vaccine refusal and dropout[23]. It’s crucial to educate parents about the typical and mild nature of post-vaccination reactions and provide reassurance from healthcare providers to mitigate fears and ensure continued adherence to the vaccine. schedule.

Similar recommendations have been made in studies from other low- and middle-income countries (LMICs), where improving vaccine service delivery and reducing logistical barriers significantly increased vaccination rates[24].

Furthermore, integrating mobile vaccination units and community-based healthcare approaches may help overcome the logistical challenges faced by families with multiple children or those living far from healthcare facilities. This approach has been successfully implemented in various countries, including India and Nigeria, where mobile clinics helped to improve vaccine coverage in remote and underserved areas[25]

## Conclusion

Bangladesh aims to eliminate measles by 2026, targeting at least 95% vaccination coverage. This study identified key socio-demographic and service-related factors associated with measles vaccine dropout in urban slums. Children of homemaker mothers, those with higher birth order, and families experiencing long waiting times at vaccination sites were more likely to miss the measles vaccination schedule. Targeted interventions such as reducing service delays, enhancing community outreach, and improving parental awareness are essential to increase vaccine adherence. Implementing these evidence-based strategies can strengthen Bangladesh’s immunization program and advance progress toward measles elimination.

## Data Availability

The datasets generated and/or analyzed during the current study are stored at the Institute of Epidemiology, Disease Control and Research (IEDCR). Data can be made available upon reasonable request by contacting IEDCR at.

## Acknowledgements

I extend my gratitude to Professor Mahmudur Rahman, MPHM, PhD, Country Director, EMPHNET, and Chair of the National Verification Committee of Measles Elimination in Bangladesh, for his technical and academic support. I also thank Dr. Mahbubur Rahman, Course Coordinator, FETP, B, for his continuous mentoring and inspiration. Finally, I acknowledge the authority and staff of Dhaka North City Corporation for their cooperation and assistance during this study.

